# Susceptibility to Residual Inhibition is Associated with Hearing Loss and Tinnitus Chronicity

**DOI:** 10.1101/2020.08.05.20167346

**Authors:** Suyi Hu, Lukas Anschuetz, Deborah A. Hall, Marco Caversaccio, Wilhelm Wimmer

## Abstract

Residual inhibition, i.e. the temporary suppression of tinnitus loudness after acoustic stimulation, is a frequently observed phenomenon that may have prognostic value for clinical applications. However, it is unclear in which subjects residual inhibition is more likely and how stable the suppression can be induced repeatedly. The primary aim of this work was to evaluate the effect of hearing loss and tinnitus chronicity on residual inhibition susceptibility. The secondary aim was to investigate the short-term repeatability of residual inhibition. Residual inhibition was assessed in 74 tinnitus subjects with 60-second narrowband noise stimuli in 10 consecutive trials. The subjects were assigned to groups according to their depth of suppression (substantial residual inhibition vs. comparator group). In addition, a categorization in normal hearing and hearing loss groups, related to the degree of hearing loss at the frequency corresponding to the tinnitus pitch, was made. Logistic regression was used to identify factors associated with susceptibility to residual inhibition. Repeatability of residual inhibition was assessed using mixed-effects ordinal regression including post-stimulus time and repetitions as factors. Tinnitus chronicity was not associated with residual inhibition for subjects with hearing loss, while a statistically significant negative association between tinnitus chronicity and residual inhibition susceptibility was observed in normal hearing subjects (odds ratio: 0.63; CI: 0.41 to 0.83; p = 0.0076). Moreover, repeated states of suppression can be stably induced. Our results suggest that long chronicity and residual inhibition susceptibility could be indicators for hidden lesions along the auditory pathway in subjects with normal hearing thresholds at their tinnitus frequency.

## 1 Introduction

Residual inhibition (RI) refers to the phenomenon of transient tinnitus loudness suppression after exposure to an acoustic stimulus (Terry et al., 1983; Roberts et al., 2008). It was first described more than 100 years ago by Spaulding (1903) and systematically studied by Feldmann in the 1970s (Feldmann, 1971). The prevalence of RI is estimated to be over 75% in subjects with tinnitus (Vernon and Meikle, 2003; Roberts et al., 2006). In the remaining tinnitus subjects, exposure to acoustic stimuli either does not alter tinnitus perception or, in rare cases, temporarily increases tinnitus loudness (residual excitation, RE) (Sedley et al., 2012). RI can be induced by various types of stimuli, including pure-tones (Terry et al., 1983), broadband noise (Vernon and Meikle, 2003), narrow-band noise (Roberts et al., 2008) and amplitude modulated sounds (Reavis et al., 2012). It has been observed that both the duration and depth of RI (i.e., the degree of tinnitus loudness change) correlate with the intensity and spectrum of the acoustic stimulus. Notably, the maximum RI time increases nonlinearly as the duration of the stimulation gets longer (Terry et al., 1983). In the majority of subjects the suppression can last from a few seconds to minutes (Vernon and Meikle, 2003) and, in rare cases even up to several hours (Vernon, 1981; Olsen et al., 1996). RI has potential as a useful tool in clinic, notably as a diagnostic marker for subtyping and also as a prognostic indicator for individual responses to therapeutic acoustic stimulation. For example, the varying depth and duration of RI in individuals could enable a more refined tinnitus classification. In cases where positive RI leads to a transient tinnitus reduction, the procedure can also be used to reassure patients during the counseling process (Fournier et al., 2018).

Despite these potential clinical benefits, RI is under-represented in the routine assessment of tinnitus patients in clinics. A reason why could be due to uncertainties in the mechanisms underlying RI, combined with the relatively long testing times. A hypothesis has been put forward that RI is a temporary reduction of hyperactive spontaneous activity or desynchronization of excessive synchronous activity at or below the level of the auditory cortex in deafferent regions caused by hearing loss (Galazyuk et al., 2017; Sedley et al., 2012; Kahlbrock and Weisz, 2008; Roberts et al., 2008). The suppression of spontaneous activity in the inferior colliculus was reported during RI in animal experiments (Galazyuk et al., 2017). Neuroimaging studies in human subjects showed decreased delta, theta and gamma oscillations of the auditory cortex during RI, indicating a restoration of the balance between excitatory and inhibitory neural processes (Kahlbrock and Weisz, 2008; Sedley et al., 2012; Roberts et al., 2015; Adamchic et al., 2017). However, no change in delta and theta and a decrease in gamma oscillations was observed in the subjects during RE, suggesting a more complex mechanism behind RI and tinnitus (Sedley et al., 2012). Additionally, Galazyuk et al. (2017) showed in an animal study that repeated exposures to the acoustic stimulus are associated with a gradual reduction of inferior colliculus activity. This indicates a possible habituation effect of RI that might reduce the potential benefits of RI during the counseling process implying the importance of analyzing repeatability of RI using human subjects.

As part of a neuroimaging study applying RI to modulate tinnitus perception (Hu et al., 2019), we wanted to identify which factors, in addition to those already known (i.e. form, frequency, intensity and duration of the acoustic stimulus), could have a positive influence on the susceptibility of subjects to experiencing RI. Hearing loss is one of the major factors associated with tinnitus (Shargorodsky et al., 2010). Previous studies have demonstrated that the use of stimuli targeting the hearing loss frequency (which often coincides with the tinnitus frequency spectrum) was most effective for RI (Terry et al., 1983; Vernon and Meikle, 2003; Fournier et al., 2018). However, considering the presence of tinnitus in subjects with normal audiograms (Savastano, 2008), additional factors need to be considered. Sedley et al. (2016) suggested that the persistence of tinnitus is caused by changing the default prediction for silence after a certain time of tinnitus onset. Furthermore, they proposed that RI could be associated with either the change of the spontaneous activity along the auditory pathway or of the default prediction for silence. This indicates that using acoustic stimuli targeting deafferent regions caused by hearing loss might influence RI susceptibility. We hypothesized that subjects with hearing loss accompanied by abnormal spontaneous activity along the auditory pathway may be overall more susceptible to RI. Moreover, we hypothesized that for subjects without hearing loss, tinnitus chronicity may be influential to RI susceptibility due to a change in default prediction. Therefore, the primary aim of this study was to evaluate RI susceptibility under consideration of a hearing loss category and tinnitus chronicity. The secondary aim was to investigate whether RI can be repeatedly induced after 10 repetitions in a short-term setting.

## Methods

### Study Design and Setting

The presented analysis was performed using the screening data collected in an ongoing study being conducted at the Bern University Hospital, Inselspital, Bern, Switzerland (Hu et al., 2019). The study was approved by the cantonal ethics committee of Bern, Switzerland (reference number: KEK-BE 2017-02037). The participants were recruited via the outpatient clinic in our department. All participants gave written informed consent about the usage of their data before starting the screening stage. Data of the period from February 1st 2018 to February 29th 2020 was used for the analysis.

### Tinnitus Subjects

The screening data of subjects meeting the following criteria were included in the analysis: (1) age ≥ 18 years; (2) subjective tinnitus that is not fluctuating; (3) single-pitched tinnitus, either perceived unilaterally, bilaterally (in both ears) or centrally (in the head); (4) no “catastrophic” tinnitus, i.e. a tinnitus handicap inventory (THI) score less than 76 (Newman et al., 1996), (5) no change of tinnitus form (pure-tone or noise-like) or pitch after RI stimulus presentation, and (6) no enhancement of tinnitus loudness (Residual Excitation) after exposure to an acoustic stimulus. Data from subjects with bilateral tinnitus experiencing different levels of tinnitus suppression in each ear were excluded from the analysis.

### Audiometry and Tinnitus Assessment

For a detailed description of the assessment procedure and measurement setup please refer to the protocol of the accompanying study (Hu et al., 2019). As part of the screening procedure, all participants completed a questionnaire containing information on the patients’ medical history, age (in years) and tinnitus chronicity (in years), the THI questionnaire and the Hospital Anxiety and Depression Scale (HADS) questionnaire (Zigmond and Snaith, 1983).

All psychoacoustic measurements were performed inside an acoustic chamber. To generate the acoustic stimuli, we used a custom-written Matlab script (The MathWorks Inc, v.2017b) with the Psychophysics-Toolbox extension (Brainard, 1997). The stimuli were presented through an external sound card (Scarlett2i2, FocusRite) and high-definition in-ear headphones (E1001, Triple-Driver, 1MORE Inc). Calibration of the stimuli was performed using a head and torso simulator, including 2 ear simulators (Type 4128, Brüel & Kjaer) and an audio analyzer (UPV Audio analyzer DC-250 kHz, Rohde & Schwarz). For the measurement of air conduction hearing thresholds (in dB sound pressure level, SPL) an extended pure-tone audiometry was performed at 0.125, 0.25, 0.5, 1, 2, 3, 4, 6, 8, 9, 10, 11, 12 and 13 kHz. The subjects also reported their tinnitus laterality (i.e. unilateral left, unilateral right, bilateral “in both ears” or central “in the head”) and form (i.e. tonal or noise-like). Tinnitus pitch (in kHz) and loudness (in dB SPL) were estimated by matching with an ipsilateral stimulus in the range of 0.125 to 13 kHz, using either pure-tone or third-octave band noise stimuli, depending on the tinnitus form indicated.

### Residual Inhibition Assessment

For RI assessment, we used a 60-second third-octave band noise stimulus, whose center frequency was set to the tinnitus pitch. For improved comparability, we additionally measured the air conduction threshold (in dB SPL), minimum masking level (MML; in dB SPL) and loudness discomfort level (LDL; in dB SPL) using the RI stimulus. In case of unilateral tinnitus, the RI stimulus was presented ipsilaterally at a level 20 dB above the MML. Contralaterally, the stimulation level was adjusted so that it was at the same sensation level (SL) as the ipsilateral stimulus. This was achieved by adding the difference between the RI stimulus level of the tinnitus ear and the ipsilateral third-octave narrow band noise threshold to the third-octave narrow band noise threshold of the contralateral ear. In case of a bilateral or central tinnitus, both ears were stimulated with the same stimulus 20 dB above the MML. To assess the short-term repeatability of RI, subjects who reported suppression of their tinnitus after acoustic stimulation were repeatedly examined in 10 consecutive trials. Between the individual repetitions, the subjects used a response box to rate the change in tinnitus loudness on an 11-point Likert scale (range: −5 to 5; −5 complete suppression, 0 no change, +5 enhancement) until it returned to its previous level. To assess the time-related change in RI depth, the time of each rating was recorded (denoted “RI time”). After the tinnitus loudness had returned to its baseline level, the next repetition was initiated. Our primary outcome measure of RI likelihood was the maximum RI depth after stimulus offset averaged over the 10 repetitions. Subjects who achieved an averaged maximum RI depth of −5 or −4 (corresponding to a complete or almost complete suppression of tinnitus) were assigned to the “RI group” (i.e., having RI capability), while the remaining subjects were assigned to the “Comparator” group (no substantial suppression). The conservative threshold of −4 was chosen based on the assumption that substantial RI should be observed in the subjects when using a RI stimulation level of 20 dB above MML. The time after which the tinnitus returned to the loudness before the stimulus (i.e. the subject presses 0 after RI) was defined as maximum RI time (in seconds). Only data of subjects from the RI group were included in the analysis of the short-term repeatability of RI. For analysis, the RI depth and RI time of all repetition trials were used as secondary outcome measures.

### Statistical Analysis

Descriptive statistics were used to summarize demographic data, tinnitus characteristics and RI outcomes. The hearing thresholds were converted from dB SPL to dB hearing level (HL) using the reference values specified in the literature for pure-tone (reference age group: 10-21 years (Lee et al., 2012)). For the hearing loss categorization, the averaged hearing threshold at the frequency corresponding to the tinnitus pitch and the two adjacent frequencies of the tinnitus ear was used (hearing loss: > 25 dB HL; normal hearing ≤ 25 dB HL). On average, the normal hearing group subjects were 25.0 years younger than the subjects in the hearing loss group (CI: 20.0, 29.5; p < 0.001). In addition, we calculated the Pearson correlation coefficients between the HADS and THI questionnaires, which are known to be correlated (Andersson et al., 2009). This was confirmed by the correlation coefficients of 0.58 between THI and HADS-A responses (CI: 0.37 to 0.73; p < 0.001), 0.57 between THI and HADS-D responses (CI: 0.39 to 0.70; p < 0.001) and 0.66 between HADS-A and HADS-D responses (CI: 0.48 to 0.80; p< 0.001).

To test the differences between the RI and Comparator groups for demographic and tinnitus characteristics, we applied the Mann-Whitney-U and *χ*^2^ tests for continuous and categorical variables, respectively. We used multivariable logistic regression to compute the odds ratios (ORs) for the susceptibility to substantial RI (i.e. almost complete or complete RI), with the dependent outcome variable defined as the RI group (Comparator vs RI). The initial model was populated with effects for hearing loss category, age, gender, tinnitus form, tinnitus laterality, tinnitus chronicity, THI score, tinnitus pitch, MML, and LDL. The HADS scores were not included because of the strong collinearity with THI scores. An interaction between hearing category and tinnitus chronicity was included to model dependencies between the variables. A step-wise backward elimination based on Akaike’s Information Criterion was applied for model selection, resulting a final model that consisted of hearing category, tinnitus chronicity and the interaction term between both variables.

The short-term repeatability of RI was assessed using an ordinal mixed-effects model with RI depth (i.e., levels −5 to 0) as the ordinal dependent outcome. The variables RI time (time after stimulus offset) and repetition (trials 1 to 10) were included as fixed effects with an interaction. All other covariates showed a lack of statistical significance. The subject identity number was included as random intercept to account for repeated-measures. All statistics were performed using the R environment (version 3.6.2) (R Core Team, 2017), with the modules to “mixor” (Archer et al., 2018) for ordinal mixed-effects model fitting.

## Results

### Data Characteristics

From the data set of 109 screened tinnitus subjects, the records of 74 subjects were included in the analysis (Figure 1). A summary of the data is given in Table 1. The majority of the subjects indicated a pure-tone tinnitus (78%). Interestingly, almost 3/4 of the subjects experienced their tinnitus pitch at a frequency above 8 kHz (average tinnitus pitch of 9.2 kHz), i.e. within a test range usually not covered in routine clinical audiometry. The mean reported tinnitus loudness was 7.2 dB SL. The THI and HADS scores indicated slight tinnitus severity and low levels of anxiety and depression of the subjects.

The proportions of subjects assigned to the “RI” and “Comparator” groups were 65% and 35%, respectively. Two of the 48 subjects in the RI group experienced long-term RI (maximum RI time ≥ 5 minutes). Since the maximum RI time could not be measured within the time available in the screening session, the 2 subjects were excluded from the descriptive statistics for the maximum RI time.

**Figure 1.**
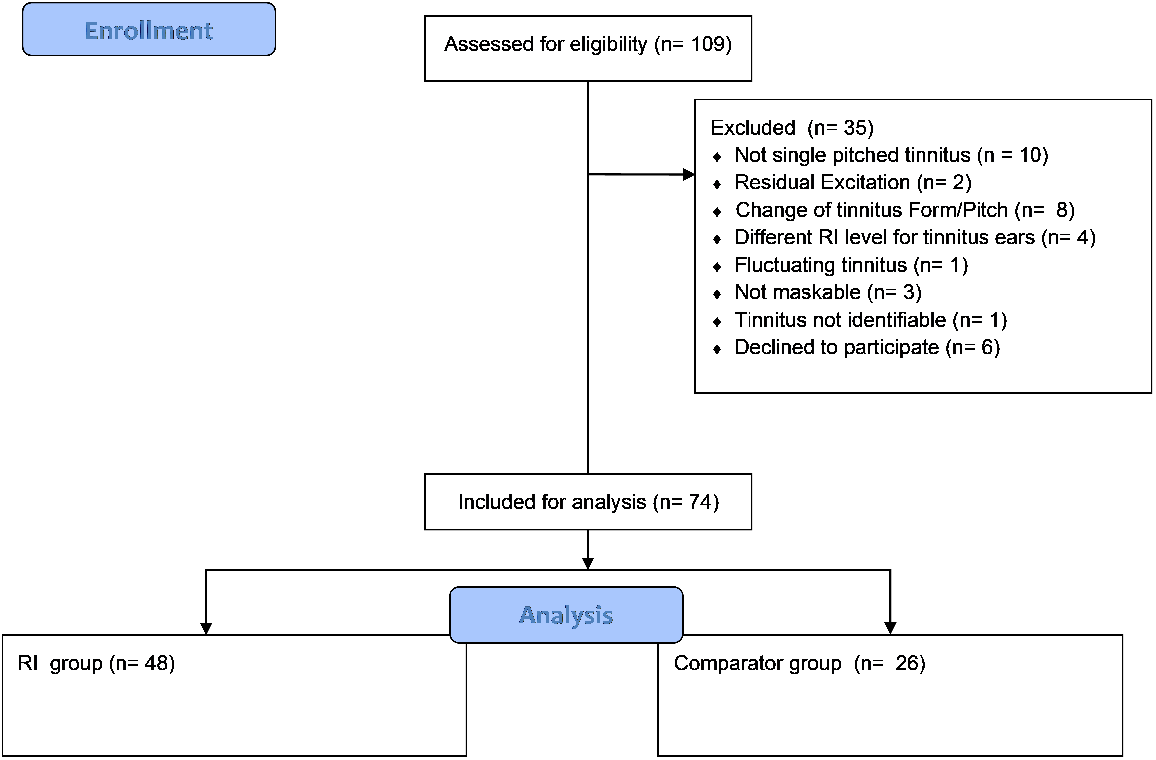
Flowchart for screening data inclusion. RI = residual inhibition.

**Table 1.** Overview of demographic details, tinnitus characteristics and residual inhibition outcomes. Comparator = subjects with no substantial residual inhibition (RI depth > −4); RI = subjects with (almost) complete residual inhibition (RI depth ≤ −4); HL = hearing level; PTA = pure-tone average over 0.5, 1, 2, 4 and 8 kHz; THI = tinnitus handicap inventory; HADS = hospital anxiety and depression scale; SL = sensation level. Continuous variables are summarized with their mean values (± standard deviation).

|  | Comparator (n=26) | RI (n=48) | All (n=74) |
| --- | --- | --- | --- |
| **Hearing category** |  |  |  |
| Hearing loss group | 12 (46%) | 39 (81%) | 51 (69%) |
| Normal hearing group | 14 (54%) | 9 (19%) | 23 (31%) |
| **Gender** |  |  |  |
| Female | 9 (35%) | 16 (33%) | 25 (34%) |
| Male | 17 (65%) | 32 (67%) | 49 (66%) |
| Age, years | 41.8 ($\pm 16.1$) | 49.7 ($\pm 14.1$) | 46.9 ($\pm 15.2$) |
| Hearing threshold at tinnitus pitch, dB HL | 33.5 ($\pm 29.9$) | 44.4 ($\pm 27.3$) | 40.5 ($\pm 28.5$) |
| Hearing threshold (PTA), dB HL | 28.7 ($\pm 13.6$) | 32.6 ($\pm 15.3$) | 31.2 ($\pm 14.7$) |
| **Tinnitus chronicity, years** | 8.9 ($\pm 7.0$) | 10.0 ($\pm 10.2$) | 9.6 ($\pm 9.2$) |
| **Tinnitus form** |  |  |  |
| Noise-like | 7 (27%) | 9 (19%) | 16 (22%) |
| Pure-tone | 19 (73%) | 39 (81%) | 58 (78%) |
| **Tinnitus laterality** |  |  |  |
| Bilateral | 16 (62%) | 30 (62%) | 46 (62%) |
| Unilateral | 10 (38%) | 18 (38%) | 28 (38%) |
| Tinnitus pitch, kHz | 10.0 ($\pm 2.1$) | 8.7 ($\pm 3.1$) | 9.2 ($\pm 2.8$) |
| Tinnitus loudness, dB SL | 7.2 ($\pm 7.9$) | 7.3 ($\pm 9.0$) | 7.2 ($\pm 8.6$) |
| Minimum masking level, dB SL | 16.5 ($\pm 12.0$) | 16.6 ($\pm 12.3$) | 16.6 ($\pm 12.1$) |
| Loudness discomfort level, dB SL | 47.1 ($\pm 14.7$) | 45.4 ($\pm 15.7$) | 46.0 ($\pm 15.3$) |
| THI score | 28.7 ($\pm 20.3$) | 28.8 ($\pm 20.3$) | 28.8 ($\pm 20.2$) |
| HADS-A score | 4.7 ($\pm 3.8$) | 5.2 ($\pm 3.1$) | 5.0 ($\pm 3.3$) |
| HADS-D score | 3.1 ($\pm 3.3$) | 3.8 ($\pm 3.3$) | 3.5 ($\pm 3.3$) |
| **Averaged maximum RI depth** | -1.3 ($\pm 1.6$) | -4.8 ($\pm 0.3$) | -3.5 ($\pm 2.0$) |
| **Averaged maximum RI time, seconds** | 21.8 ($\pm 29.1$) | 93.3 ($\pm 49.4$) | 67.5 ($\pm 55.1$) |

### Susceptibility to Residual Inhibition

The demographic results from Table 1 showed a higher percentage of “Comparator” subjects in the group with normal hearing (14 out of 23) than in the group with hearing loss (12 out of 51). The comparison between “RI” and “Comparator” in different hearing categories calculated with *χ*^2^ tests revealed a statistically significant difference (p = 0.0043) indicating that subjects with hearing loss at their tinnitus frequency are more susceptible to RI. Additionally, with the exception of age, which showed a trend toward younger subjects in the Comparator group (age difference of −8.5 years, CI: −18.1 to −0.02; p = 0.049), no statistically significant differences between the groups were observed in the other characteristics tested.

The results of the logistic regression analysis for the RI susceptibility are presented in Table 2. Tinnitus chronicity for hearing loss group did not have a statistically significant effect on RI susceptibility. However, statistically significant ORs for tinnitus chronicity for normal hearing group were observed. For a 1-year increment in tinnitus chronicity, the probability for RI susceptibility decreased by a factor of 0.63 (CI; 0.41, 0.83; p = 0.0076). These results suggest that tinnitus chronicity only affects the RI susceptibility for subjects with hearing thresholds at their tinnitus frequency ≤ 25 dB HL. Moreover, subjects with shorter tinnitus chronicity are more susceptible to RI.

**Table 2.** Logistic regression odds ratios with respect to the “Comparator” group (i.e. no substantial residual inhibition).

|  | Odds ratio | Confidence interval |  | P value |
| --- | --- | --- | --- | --- |
|  |  | 2.5% | 97.5% |  |
| (Intercept) | 2.21 | 0.85 | 6.27 | 0.12 |
| **Hearing category** (normal hearing) | 2.86 | 0.40 | 32.27 | 0.33 |
| **Tinnitus chronicity** | 1.07 | 0.98 | 1.23 | 0.19 |
| **Hearing category** (normal hearing) : **tinnitus chronicity** | 0.63 | 0.41 | 0.83 | 0.0076 |

### Short-term Repeatability of Residual Inhibition

In general, the RI depth and course of recovery were stable over the 10 repetitions for each individual. Statistically significant effects were observed for RI time, repetition and their interaction term (see Table 3). Obviously, the chance for stronger suppression decreases after stimulus offset (i.e., for longer RI times). Figure 2 illustrates the probability of reaching the different RI depth levels (−5 to 0) for the first repetition as a function of RI time. Approximately 100 seconds after stimulation offset, the majority of subjects will either perceive their tinnitus with a slight suppression (RI depth level −1) or its initial loudness (RI depth level 0). Moreover, the more repetitions are performed, the higher the probability to experience complete RI (i.e. an RI depth level of −5). Figure 3 illustrates the effect of the interaction term between RI time and repetition. After 10 repetitions, the probability of a maximum RI depth of −5 increases, while the maximum RI time (i.e. return to RI depth 0) occurs slightly earlier. This suggests that with the given conditions used during our assessment (i.e. 60 seconds stimulus, 10 repetitions, stimulus level at MML +20 dB) stable repeated RI phenomena can be generated.

**Figure 2.**
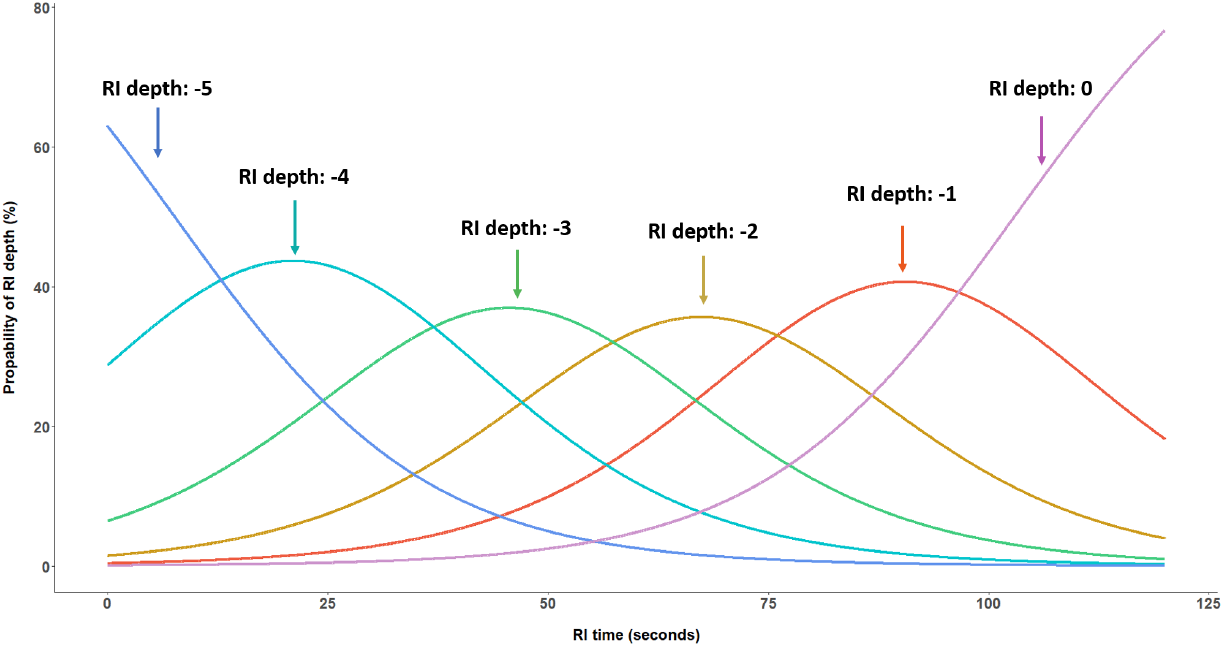
Probability of experiencing a residual inhibition (RI) depth level between −5 (complete suppression of tinnitus) and 0 (return of tinnitus loudness to the initial level) after stimulus offset for the 1^st^ repetition. Subjects with substantial RI (n=48) were included in the analysis.

**Figure 3.**
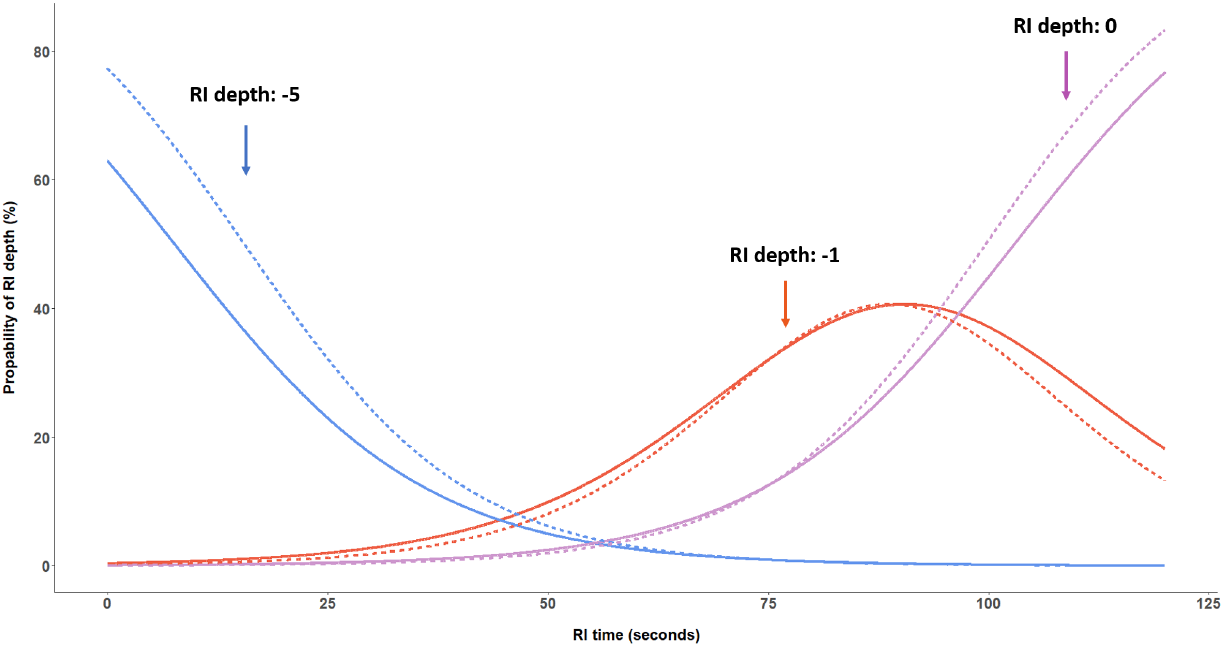
Probability of experiencing residual inhibition (RI) depth levels of −5 (complete suppression), −1 (weak suppression) and 0 (return to initial tinnitus loudness) after stimulus offset for the 1^st^ (solid lines) and the 10^th^ repetition (dashed lines).

**Table 3.** Mixed-effects ordinal regression odds ratios with respect to the RI depth level 0 (return of tinnitus loudness to the pre-stimulus level).

|  | Odds ratio | Confidence interval |  | P value |
| --- | --- | --- | --- | --- |
|  |  | 2.5% | 97.5% |  |
| (Intercept) | 1.57 | 0.77 | 3.22 | 0.21 |
| **RI depth** |  |  |  |  |
| level -4 | 6.50 | 5.85 | 7.22 | < 0.001 |
| level -3 | 30.63 | 27.18 | 34.52 | < 0.001 |
| level -2 | 135.83 | 117.18 | 157.45 | < 0.001 |
| level -1 | 762.38 | 682.08 | 852.14 | < 0.001 |
| **RI time** | 0.93 | 0.93 | 0.94 | < 0.001 |
| **Repetition** | 1.08 | 1.05 | 1.11 | < 0.001 |
| **RI time : repetition** | 0.9989 | 0.9986 | 0.9993 | < 0.001 |
| Subject ID (random intercept) | 10.11 | 2.28 | 44.77 | 0.0023 |

## Discussion

The main finding of our study is that tinnitus chronicity is negatively associated with RI susceptibility in subjects with normal hearing thresholds at their tinnitus frequency. In addition, the tinnitus tends to be more susceptible to transient modulation in subjects with hearing loss than in normal hearing subjects. In combination with the observed negative influence of chronicity in the normal hearing group, the difference in RI susceptibility based on hearing categorization may enable refined tinnitus subtyping. The higher susceptibility to RI in the hearing loss group suggests higher weighting of peripheral caused tinnitus, while the influence of chronicity in the normal hearing group indicates the maintenance of tinnitus in central systems. Furthermore, we found that consecutive repetitions of acoustic stimulation provide stable RI conditions. This finding validates that RI can be used to induce repeated states with or without tinnitus in the same subject, which is important in the context of within-subject comparison studies (e.g. (Hu et al., 2019)). The prevalence of RI observed in our study, i.e. 58 of 74 subjects (78%) with some degree of residual inhibition and 48 subjects (65%) with substantial tinnitus suppression, is comparable to previous studies reporting a prevalence between 61.5% and 88.0% (Deklerck et al., 2019; Vernon and Meikle, 2003; Henry et al., 2000; Roberts et al., 2008). The mean maximum RI time of 93.3 seconds is comparable to the findings of Vernon and Meikle (2003). In our cohort, 78% of the participants indicated a tinnitus pitch equal or higher than 9 kHz, emphasizing the importance of extended audiometry in the clinical routine assessment of tinnitus.

### Susceptibility to Residual Inhibition

The current assumption of the underlying mechanism is that RI is produced by neuronal changes in excessive activity at peripheral or central levels caused by hearing loss following acoustic stimulation in the deafferent regions. (Roberts et al., 2008; Fournier et al., 2018). Previous studies have shown that particular characteristics of the acoustic stimulus targeting tinnitus and hearing lesions are known to influence the depth and duration of RI. Terry et al. (1983) observed that the maximum RI time increases in a logarithmic fashion with increasing stimulus duration, eventually saturating for stimuli lasting longer than 60 seconds. Moreover, it is known that an acoustic stimulation resembling the hearing loss that often coincides with the tinnitus spectrum induces RI more effectively (Roberts et al., 2008; Fournier et al., 2018). However, despite the fact that hearing loss is one of the main factors contributing to tinnitus, it is not a necessary condition. Tinnitus subjects with normal hearing are not unusual (Savastano, 2008; Henry et al., 2008). Studies on neural imaging demonstrated abnormal activity and connectivity in and with other brain regions, suggesting involvement of other brain networks that mediate perception, distress, saliency, memory and attention (De Ridder et al., 2011; Elgoyhen et al., 2015). Furthermore, Sedley et al. (2016) proposed that the persistence of tinnitus is caused by resetting the default prediction from experiencing ‘silence’ to ‘tinnitus’ after long chronicity, which prevents spontaneous activities from being ignored as noise. Therefore, in addition to the decrease in spontaneous activities (or central gain), RI could be presented as a temporal reset of the default prediction to ‘silence’. Based on this hypothesis, we argue that the mechanism of RI might be different in subjects with normal hearing than in subjects with hearing loss who received sufficient acoustic stimulation in the deafferent regions, resulting in neural adaptation of excessive activity. Therefore, using acoustic stimulation targeting deafferent regions could be more effective for producing RI in hearing impaired subjects. This was observed in our data and a similar tendency was observed in the literature (Roberts et al., 2008; Fournier et al., 2018). RI in normal hearing subjects on the other hand, could rather be explained by a normalization of the incorrect default prediction. Since resetting of the default prediction occurs after a certain time of the tinnitus onset, we presume that the precision of the incorrect default prediction increases with chronicity and becomes less changeable.

Our findings raise an interesting point in the context of tinnitus management strategies. Normal hearing tinnitus subjects with RI susceptibility could represent a group with their default prediction being susceptible to modulation, which could indicate a higher likelihood to benefit from interventions. In addition, our data showed decreasing RI susceptibility with increasing tinnitus chronicity. We assume that hidden hearing lesions might be present in subjects with normal audiograms, longer tinnitus chronicity and RI susceptibility. However, this hypothesis requires testing in a case-controlled manner including assessments considering hair cells and postsynaptic structures function, such as otoacoustic emissions and auditory brainstem response recordings. Furthermore, subjects with hearing lesions may be more susceptible to therapeutic benefits by means of acoustic stimulation targeting the deafferent regions (i.e. use of hearing aids), while strategies aiming to normalize default brain prediction (i.e. reduction of attention to tinnitus through therapy) might be more suitable for subjects without hearing lesions.

### Short-term Repeatability of Residual Inhibition

Previous studies demonstrated that RI can be consistently reproducible between sessions indicating that there is no long-term adaptation affecting test-retest assessment (Roberts et al., 2008; Deklerck et al., 2019). However, the effect of consecutive repeated stimulation on shortterm adaptation and the robustness of RI has not yet been comprehensively studied in human subjects. The results of the ordinal mixed-effects model showed an increased probability for a reduced maximum RI time after several repetitions. In an animal study, a shortening of the suppression time of spontaneous firing rates of the inferior colliculus after consecutive stimulation (Galazyuk et al., 2017) was observed. Similarly, a study with a single human subject reported the reduction of the maximum RI duration after repeated stimulation (Sedley et al., 2015). We also observed an effect of repetitions on the RI depth, with a tendency to experience stronger suppression after more repetitions. In summary, our results suggest that the subjects in our study experienced stronger RI depths, however slightly shorter maximum RI times with an increasing number of repetitions. Nevertheless, the low magnitude of the effects suggests stable RI after repeated stimulation. Our analysis demonstrates that with the test conditions applied in our assessment procedure (i.e. 60 seconds stimulus, 10 repetitions, RI stimulus level at MML + 20 dB) stable repeated RI phenomena can be induced. In addition to its use in comparative within-subject studies, the stability of RI, with its ability to modulate tinnitus perception, indicates potential applications during the therapeutic counseling process.

Our analysis suggests the possibility that two different RI mechanisms could synergistically affect tinnitus subjects with and without hearing loss, but with different weightings. Normal hearing thresholds at the tinnitus frequency, longer chronicity and RI susceptibility could be indicative for hidden hearing lesions suggesting additional hearing assessments for these subjects might be required. By excluding hearing lesions, it is assumed that subjects with RI, indicating a weaker incorrect default prediction, could benefit more from an intervention. In addition, we demonstrated that RI robust mechanism for generating repeated states with and without tinnitus, as required for within-subject comparison studies.

## Data Availability

The datasets generated during and/or analysed during the current study are available from the corresponding author on reasonable request.

## Acknowledgements

This work was supported by the Infrastructure Grant of the University of Bern, Bern, Switzerland and Bernafon AG, Bern, Switzerland.

